# White Matter Pathway-Specific Effects of Subthalamic Deep Brain Stimulation on Vergence Motor Strategy and Binocular Stability

**DOI:** 10.64898/2026.11.14.26368127

**Authors:** Ibrahim Quagraine, Aratrik Guha, Camilla Kilbane, Fatema F Ghasia, Aasef G. Shaikh

## Abstract

**Objective:** Clear vision across viewing distances requires the eyes to achieve binocular alignment and hold it steady. Alignment can be established through smooth dysconjugate eye movement, vergence, alone or in conjunction with rapid saccades. The circuits governing movement strategy and subsequent gaze holding remain less understood.

**Methods:** We used subthalamic nucleus (STN) deep brain stimulation (DBS) to probe these components in 23 participants with Parkinson’s disease (PD), comparing binocular eye movements with stimulation on and off and relating behavioral changes to modeled stimulation anatomy.

**Results:** Combined saccade–vergence responses predominated in both stimulation states, but individual strategy changes were associated with stimulation location. Greater field of Forel coverage was associated with reduced saccade-assisted divergence, and stimulation anatomy helped predict individual changes in this movement strategy. Greater right associative STN coverage correlated with shorter convergence latency. The clearest group effect emerged after movement ended; DBS reduced gaze-holding variance by 37.0% after convergence and 29.4% after divergence, an overall reduction of 33.3% (95% confidence interval, 13.7–49.3%; p≈0.008). Participants with improved gaze holding had numerically greater modeled exposure to Forel and cerebellothalamic pathways.

**Conclusions:** These findings suggest distinct contributions of associative STN to initiation, consistent with go/no-go control, and Forel pathways to movement strategy. Improved gaze holding raises the novel possibility that stimulation engaging Forel and cerebellar outflow pathways enhances vergence neural integration.

**Significance:** Binocular eye movements reveal how basal ganglia and cerebellar pathways establish movement strategy and sustain alignment, guiding targeted stimulation studies in PD.

## Introduction

Looking up from a distant object and back to a line of print requires the eyes to change their relative angle (vergence) and hold the new alignment while taking in visual information. In Parkinson’s disease (PD), abnormal basal ganglia activity may disrupt this process, directly or indirectly, altering how and when a vergence begins (latency), whether there is a slow smooth vergence or it is associated with rapid saccades (motor strategy), and how steadily the eyes maintain their position after vergence (gaze holding) (Almer et al., 2012).

*Motor Strategy and Latency.* A shift in viewing depth can occur through vergence alone or in combination with a disconjugate saccade, rapidly reducing vergence error. Basal ganglia output and subthalamic nucleus (STN) activity may influence when a depth shift begins, whether a saccade is recruited, and when and how much it contributes. These effects can dissociate. More frequent saccades may not improve accuracy, and a larger saccadic contribution may leave final vergence gain unchanged(Busettini and Mays, 2005, Quinet et al., 2020, Zee et al., 1992).

Primate physiology identifies several routes for this control. Neurons in and around the supraoculomotor area (SOA) encode vergence velocity, angle, or both(Mays, 1984, Mays et al., 1986, Zhang et al., 1991). The STN may influence saccade initiation through the substantia nigra pars reticulata, whose inhibitory output regulates the superior colliculus(Hikosaka and Wurtz, 1985, Van Horn et al., 2013). Saccadic assistance allows different movement combinations to achieve the same fixation depth(Busettini and Mays, 2005). Collicular projections to the SOA and pontocerebellar circuits involving the nucleus reticularis tegmenti pontis (NRTP) may coordinate saccades with vergence(Bostan et al., 2010, Gamlin and Clarke, 1995, Glickstein et al., 1990, May et al., 2021, Stanton et al., 1988, Takeichi et al., 2005). Cerebellar fastigial and interposed nuclei project to the superior colliculus, midbrain near-response regions, and central mesencephalic reticular formation (cMRF)(Bohlen et al., 2021, May et al., 1990, May et al., 1992). In the cMRF, saccade– vergence burst neurons encode vergence velocity during disconjugate saccades. The saccadic contribution to a depth shift depends on the interaction between the burst and the remaining vergence error(Busettini and Mays, 2005, Quinet et al., 2020).

STN-region deep brain stimulation (DBS) provides an opportunity to probe these circuits in humans. We hypothesized that stimulation alters vergence latency and saccade recruitment through modulation of frontal–basal ganglia signaling in the associative STN and possible antidromic recruitment of nearby cerebellothalamic fibers. Altered cerebellar output could change when the saccadic burst arrives relative to the remaining vergence error and how much it contributes to binocular alignment.

*Gaze Holding and Vergence Neural Integration.* Maintaining alignment after a depth shift requires a vergence neural integrator that converts brief velocity commands into sustained position signals. The velocity and angle signals encoded in and around the SOA make this region a candidate substrate(Mays, 1984, Mays et al., 1986, Quinet et al., 2025). Fastigial and posterior interposed projections could allow the cerebellum to tune these sustained signals(Bohlen et al., 2021). Cerebellothalamic fibers near the field of Forel are accessible to STN-region DBS(Gallay et al., 2008). We therefore hypothesized that antidromic modulation of cerebellar output could improve vergence integration, producing steadier gaze holding after depth shifts.

To test these hypotheses, we compared DBS on and off within PD participants, measuring vergence latency, endpoint accuracy and absolute error, saccade recruitment and timing, and gaze stability after convergence and divergence. We then overlaid modeled stimulation fields on each participant’s MRI to determine whether engagement of subthalamic regions and nearby pathways was associated with changes in vergence measures, including latency, movement strategy, or gaze holding. This approach examined whether stimulation of different regions and pathways were associated with distinct effects on vergence movements and subsequent gaze stability.

## Methods

We studied 23 participants with PD defined by UK Brain Bank Clinical Criteria. All patients were consented to the standards of the Declaration of Helsinki. The protocol was approved by Louis Stokes Cleveland VA Medical Center and Cleveland Clinic Institutional Review Boards. The participants signed on approved informed consent form.

### Eye Movement Measurements

After clinical neurological and ophthalmological examination, all participants had objective eye movement assessments on the same day. The participants were seated in a chair with chin and forehead secured in front of the ocular tracking system independently measuring both eyes movements in horizontal and vertical directions (500 Hz sampling g frequency and 0.1 degree spatial resolution; EyeLink 1000^TM^, SR Research, Mississauga, ON); see our lab’s previous publications for details(Balta Beylergil et al., 2024, Beylergil et al., 2022, Beylergil et al., 2021, Gupta et al., 2021a, Gupta et al., 2021b, Gupta et al., 2023, Nambiar et al., 2025). To summarize, each session began with three-point calibration in the following horizontal and vertical components subtended by the binocular eye positions: 0°and 10°, 15° and 10°, 15° and 10°. In order to assess vergence, the light emitting diode (LED) targets were placed within a sagittal plane at 20, 55, 150, and 244 cm.

Convergence was measured as the participants moved their gaze to a near LED target from one farther away and then vice versa for divergence. Vergence component latency was recorded as an eye movement shift from baseline at least 0.5° with a velocity greater than 3°/s following shift in target. Vergence gain was captured as a ratio of the actual change in the vergence amplitude and desired change in the vergence amplitude (i.e., the vergence demand). Post-vergence stability was measured by means of variance of gaze holding.

### Localization of Electrical Stimulation

Computational analysis on patient-specific MR space confirmed the placement of the DBS electrodes, in the STN in such a way that critical sensorimotor aspect of the STN and key white-matter pathways around the STN are stimulated. The volume of activated tissue was measured from the STN and surrounding region using LeadDBS software(Horn and Kuhn, 2015, Horn et al., 2019, Neudorfer et al., 2023, Rodrigues et al., 2021, Xu et al., 2023). In addition to the STN, the VTA also extended into adjacent structures, including the fields of Forel, cerebellothalamic projections, pallidal and cerebellar projections to the thalamus adjacent to the fields of Forel, limbic, accessory, and sensorimotor regions of the subhthalamus, and caudal parts of the thalamus. Connectomic analysis was performed using the HCP 1065 atlas to objectively examine the VTA overlap with number of white-matter tracks, including fields of Forel, cerebellothalamic projections, rubrospinal tracts, and cortico-pontine pathways.

### Statistics

#### Outcomes analysis

We compared convergence and divergence during DBS on and off in PD participants, retaining paired visits and determining eligibility separately for each outcome and direction. Outcomes included latency, first, intermediate, and final gain, gaze holding, and movement strategy. Gain was signed relative to demand, with unity indicating the requested amplitude. Absolute gain errors were calculated per take as |gain − 1| and multiplied by demand to obtain angular errors. Subsequent correction was initial minus final error.

#### Behavioral analyses

Measurements were matched within participant, visit, direction, and nominal demand. Takes were averaged, using medians for latency, followed by equal averaging across matched demands and visits. Two-sided sign-flip tests assessed participant-level on-minus-off changes; participant bootstrap resampling provided confidence intervals. Permutation tests assessed strategy distributions, and paired slope comparisons assessed demand dependence. Trial-level mixed models adjusted for stimulation state, direction, their interaction, and demand, with participant and nested-visit random intercepts and participant-clustered intervals.

#### Anatomical associations and prediction

Ward clustering examined standardized behavioral changes, with silhouette scores and omission analyses assessing cluster structure. Anatomical associations were tested using Spearman correlations with 29,999 permutations and Benjamini–Hochberg correction. Eligible variables required at least 12 observations and sufficient variation.

Elastic-net models evaluated 42 anatomical predictors using outer leave-one-participant-out prediction and inner fivefold tuning, with preprocessing restricted to training folds. Prediction errors were compared with the training-cohort mean. Follow-up models examined associative STN coverage and convergence latency, and fasciculus subthalamicus activation and movement strategy.

## Results

### Clinical and ophthalmic characteristics

The paired DBS vergence analysis included 23 PD participants. Mean age was 68.2 ± 9.3 years, mean disease duration was 12.1 ± 6.7 years, and 22 of 23 participants were men. The mean motor score measured with Unified Parkinson’s Disease Rating Scale part 3 (UPDRS-III) was 36.3 ± 13.2 with DBS off and 14.2 ± 6.1 with DBS on. Mean daily levodopa intake was 769 ± 628 mg/day.

With DBS on, distance cover testing showed orthophoria in 19 of 23 participants (82.6%), whereas near cover testing documented exotropia or intermittent exotropia in 20 of 23 (87.0%). Visual acuity was 20/25 or better in 87.0% of left eyes and 78.3% of right eyes. Median measurable stereopsis was 100 arcsec (interquartile range, 60–100 arcsec). In participants with paired clinical convergence measurements, the mean on-minus-off change was −2.2 ± 6.9 for Royal Airforce (RAF) ruler distance (*n* = 17) and −4.5 ± 9.2 for distance to pen (*n* = 20). Table 1 depicts demographic, neurological, and ophthalmological clinical summary.

**Table 1.** Clinical and ophthalmic characteristics of the paired DBS vergence cohort.

| Characteristic | Summary |
|---|---|
| **Demographic and motor characteristics** |  |
| Age, years | 68.2 ± 9.3 (range, 52–87) |
| Sex | 22 male (95.7%), 1 female (4.3%) |
| Race, as recorded | 20 White (95.2%), 1 African American (4.8%) |
| Parkinson’s disease duration, years | 12.1 ± 6.7 (range, 4–30) |
| UPDRS-III Motor score, DBS OFF | 36.3 ± 13.2 |
| UPDRS-III Motor score, DBS ON | 14.2 ± 6.1 |
| UPDRS-III Motor score, paired ON-minus-OFF change | −22.1 ± 12.0 |
| Within-participant UPDRS-III motor score improvement, % | 58.2 ± 18.9 |
| Estimated total daily levodopa dose, mg/day | Median 600 [450–800], mean 769 ± 628 |
| **Ophthalmic and patient-reported measures** |  |
| Interpupillary distance, mm | 63.2 ± 3.9 |
| Distance orthophoria, DBS ON | 19 (82.6%) |
| Near exotropia or intermittent exotropia, DBS ON | 20 (87.0%) |
| Visual acuity of 20/25 or better, left eye | 20 (87.0%) |
| Visual acuity of 20/25 or better, right eye | 18 (78.3%) |
| Measurable stereopsis, arcsec | Median 100 [60–100] |
| 25-item Visual Function Questionnaire composite score | 84.7 ± 9.2 |
| Convergence Insufficiency Symptom Survey total score | 20.0 ± 9.5 |
| **Paired clinical convergence measurements** |  |
| RAF ruler distance | OFF 19.7 ± 7.0, ON 17.5 ± 7.2, change −2.2 ± 6.9 |
| Distance to pen | OFF 21.7 ± 9.7, ON 17.2 ± 9.8, change −4.5 ± 9.2 |

### Effects of DBS on vergence

We quantified the within-participant effect of STN DBS on vergence in 23 PD participants. We examined initiation, early and final responses, movement strategy (saccade vergence combination), and post-vergence gaze holding (Fig 1A-F).

**Figure 1:**
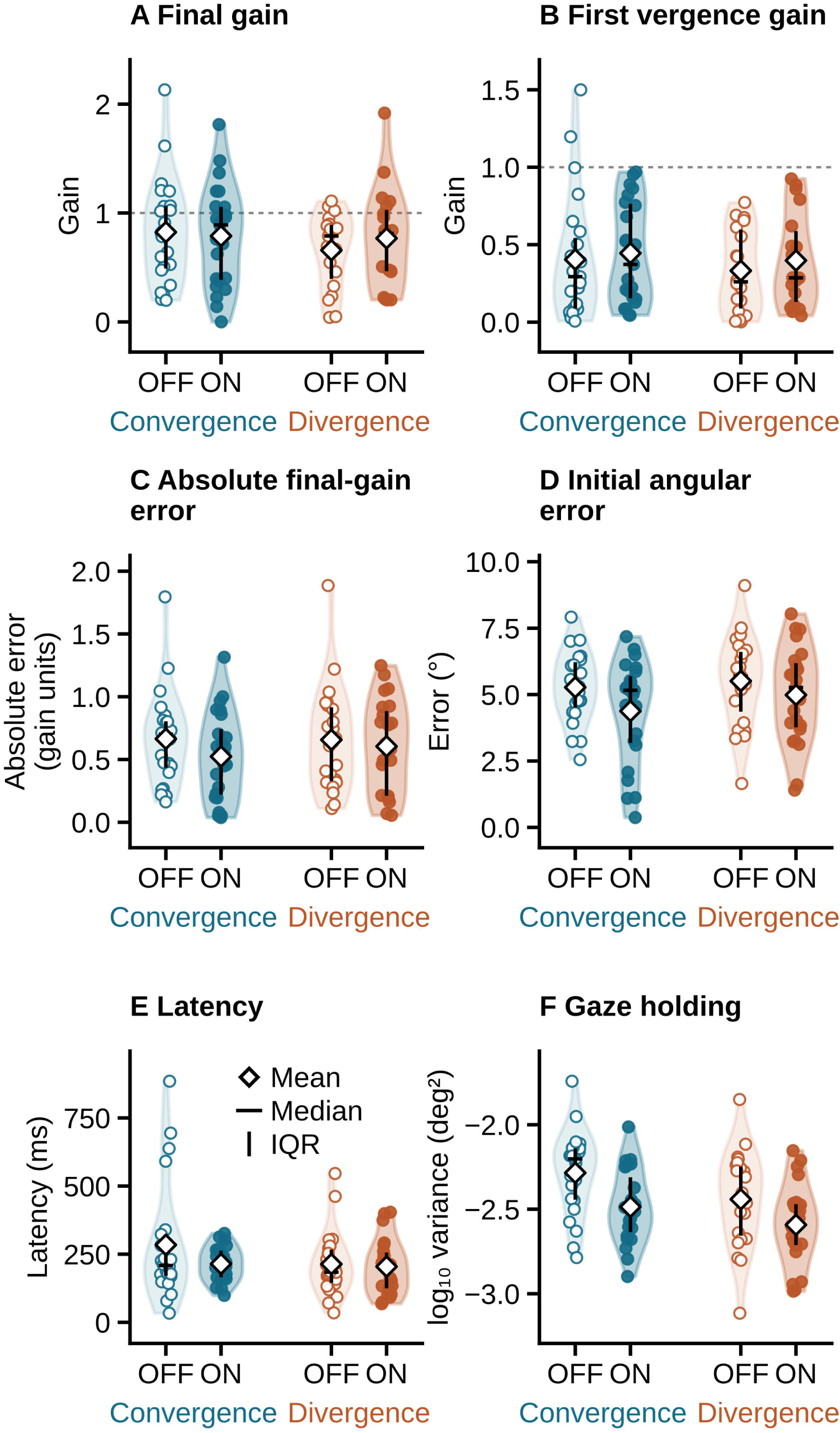
Participant-weighted distributions of six vergence measures during DBS OFF and ON. Panels A–F show final gain, first vergence gain, absolute final-gain error, initial angular error, latency, and gaze holding. Each panel contains convergence OFF/ON and divergence OFF/ON pairs. Violins show kernel density estimates with participant values overlaid; no connector lines are drawn. Open circles indicate OFF and filled circles ON; blue indicates convergence and orange divergence. White diamonds mark means; black bars show medians and interquartile ranges. Dashed lines in gain panels indicate unit gain. User-specified cutoffs were applied to participant-level state summaries: latency >1500 ms, initial angular error >15 degrees, and final or first vergence gain <0. If either state met a cutoff, both OFF and ON values were excluded from that parameter and direction, retaining paired comparisons. These are parameter-specific exclusions; the participant remains in other panels if eligible. Paired sample sizes are provided in the accompanying descriptive statistics, not printed in the panels. The mean, median and IQR key is inside panel E. Gaze holding remains on the recorded log10 variance scale. These exclusions differ from those used for the earlier clustering figures.

### DBS reduced variability during post-vergence gaze holding

Participant-weighted averages showed that STN DBS improved gaze holding in convergence as well as divergence positions (Table 2). Variance, the measure of gaze holding, decreased from 0.005 to 0.003 during convergence (37.0% reduction, paired p = 0.007, Fig 1F) and from 0.004 to 0.002 during divergence (29.4% reduction, paired p = 0.028, Fig 1F). Across the 23 participants with data in both directions, the overall reduction was 33.3% with equal weighting of convergence and divergence (participant-bootstrap 95% CI, 13.7%–49.3% reduction, paired p ≈ 0.008, Table 2).

**Table 2.** Group-level effects of DBS on vergence.

| **Measure** | **Convergence OFF → ON** | **Change** | **Paired p** | **Divergence OFF → ON** | **Change** | **Paired p** |
|---|---|---|---|---|---|---|
| Final gain | 0.757 → 0.757 | +0.001 | .99 | 0.561 → 0.625 | +0.064 | .40 |
| First vergence gain | 0.354 → 0.450 | +0.096 | .26 | 0.254 → 0.299 | +0.044 | .47 |
| Intermediate gain | 0.544 → 0.589 | +0.044 | .63 | 0.338 → 0.370 | +0.032 | .62 |
| Absolute final-gain error | 0.665 → 0.524 | −0.141 | .075 | 0.657 → 0.605 | −0.052 | .59 |
| Initial angular error, ° | 5.84 → 4.45 | −1.39 | see text | 5.51 → 4.99 | −0.52 | see text |
| Latency, ms | 378 → 286 | −92 | .52 | 280 → 204 | −76 | .38 |
| Gaze Holding | −2.285 → −2.486 | −0.201 | .007 | −2.441 → −2.592 | −0.151 | .028 |
| Vergence demand, ° | 6.899 → 6.870 | −0.028 | .38 | 6.747 → 6.721 | −0.026 | .50 |

### Final gain, first gain, and latency showed no clear average group-level change

Vergence responses often comprised a vergence movement and an accompanying disconjugate saccade, with either component occurring first. First vergence gain describes the initial vergence component, and final gain describes the alignment reached after the depth shift. First vergence gain increased in both directions (convergence, +0.10, paired p = 0.26; divergence, +0.04, paired p = 0.47, Fig 1B). The final gain was similar between stimulation states for convergence (paired p = 0.99) and increased for divergence (0.56 to 0.63, paired p = 0.40, Fig 1A).

Mean latency was shorter with DBS on in both directions, with uncertain paired estimates (Table 2). Convergence latency changed from 378 to 286 ms (−92 ms, paired p = 0.52, Fig 1E), and divergence latency from 280 to 204 ms (−76 ms, paired p = 0.38, Fig 1E).

### DBS reduced initial convergence error

To assess the accuracy of the initial vergence response, we calculated initial angular error for each trial as the absolute difference between the desired vergence movement and the amplitude of the first vergence movement. This measure depicts how far the first movement landed from the target, regardless of whether it undershot or overshot. At the group level, STN DBS significantly reduced convergence initial angular error (DBS off 5.84°, DBS on: 4.45°, Fig 1C). For divergence, the reduction in initial angular error was smaller (Off: 5.51°; On:4.99°, Fig 1C), suggesting a weaker improvement in early divergence accuracy than in convergence (Table 2). Endpoint accuracy was quantified as absolute final-gain error. During convergence, this error decreased from 0.665 to 0.524 (−0.141, paired p = 0.07, Fig 1D). The paired comparison for divergence gave p = 0.59 (Table 2).

### Strategy distributions showed no clear group-level DBS effect

Depth shifts often included a disconjugate saccade in addition to vergence. The combined response remained the predominant strategy in both DBS states, accounting for 84.3% and 84.6% of convergence responses, 83.0% and 82.2% of divergence responses, and 85.5% and 84.2% overall (OFF and ON, respectively; convergence p = 0.9, divergence p = 0.8, Table 3).

**Table 3.**
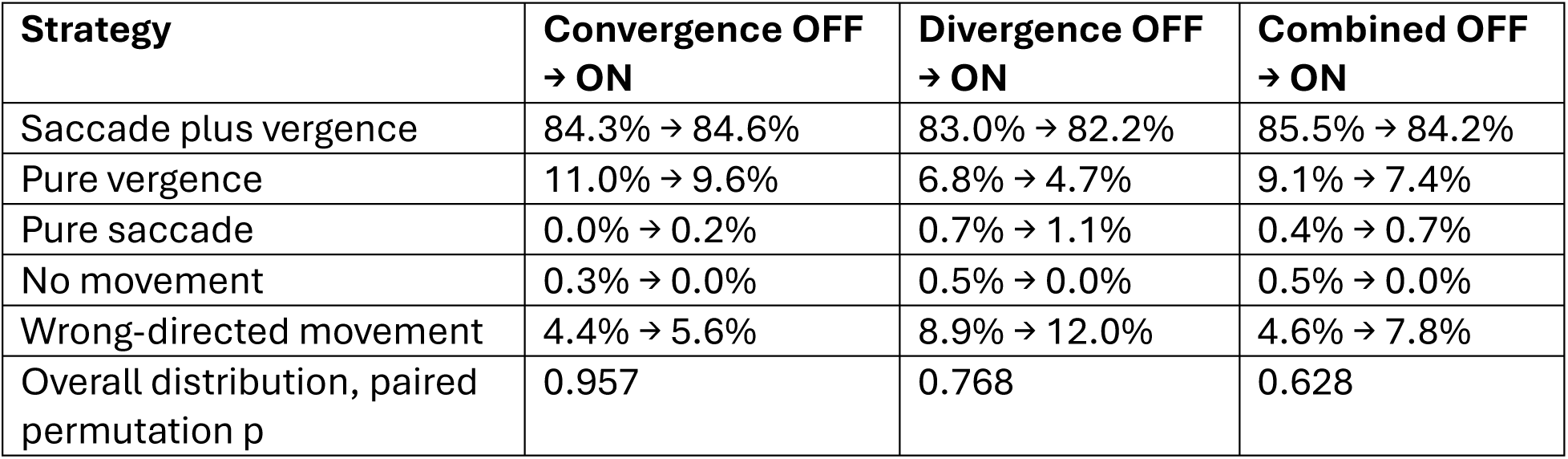
DBS effects on vergence strategy using the merged five-category classification.

### DBS effects did not differ significantly between convergence and divergence

Within-participant comparisons of DBS on–off changes showed no significant interaction with movement direction for any primary outcome (Table 4). The strongest evidence for an interaction was for final gain (convergence minus divergence, −0.10, p = 0.06). Interaction p values were 0.96 for first gain, 0.38 for absolute final-gain error, 0.46 for latency, 0.36 for gaze holding, and 0.71 for movement strategy distribution.

**Table 4.** Direction-specific comparison of DBS effects on convergence versus divergence.

| **Outcome** | **DBS change in convergence minus DBS change in divergence** | **Interaction p** |
|---|---|---|
| Final gain | −0.099 | 0.069 |
| First gain | −0.003 | 0.96 |
| Intermediate gain | −0.046 | 0.51 |
| Absolute final-gain error | −0.059 | 0.38 |
| Initial angular error, ° | larger decrease in convergence than divergence | not significant |
| Latency | +82 ms | 0.46 |
| Holding | −0.050 | 0.36 |
| Overall strategy distribution (six-category) | — | 0.71 |
| Overall strategy distribution (five-category) | — | .984 |

### DBS did not significantly alter dependence of vergence behavior on target demand

To test whether DBS altered the scaling of vergence with stimulus amplitude, we compared off-state and on-state slopes across matched low, intermediate, and high demands within each participant. Across participants, DBS did not significantly change the demand dependence of final gain, first gain, or latency. For convergence, the mean slope change was +0.021 gain units/degree for final gain (paired p = 0.32), +0.020 gain units/degree for first gain (paired p = 0.32), and −34 ms/degree for latency (paired p = 0.44). For divergence, corresponding changes were +0.012 gain units/degree (paired p = 0.41), +0.016 gain units/degree (paired p = 0.21), and +2 ms/degree (paired p = 0.95). The available data therefore did not establish that DBS altered the demand dependence of gain or latency.

### Group averages concealed heterogeneous individual responses

The small average on-off changes in gain, final accuracy, and strategy reflected cancellation among markedly different individual responses. In several participants, changes in gain magnitude, endpoint accuracy, timing, early accuracy, and movement strategy moved independently, emphasizing that these dimensions should not be conflated. For example, PD19 showed one of the clearest beneficial responses: final gain increased by 0.35 during convergence and 0.80 during divergence, while absolute final-gain error decreased by 0.85 and 0.50, respectively, and latency shortened in both directions. In contrast, PD29 and PD31 became substantially more accurate in both directions despite decreases in convergence gain (−0.73 and −0.65, respectively), consistent with correction of excessive off-state responses. At the opposite extreme, PD18 showed large increases in final gain (+1.31 during convergence and +1.05 during divergence) but became less accurate, with absolute final-gain error increasing by 0.58 and 0.84. Thus, increased gain could accompany either improvement or deterioration in endpoint performance, depending on the participant’s baseline state.

The initial-angular-error analysis reinforced this point. DBS-related increases in first movement magnitude did not necessarily imply better early accuracy; rather, the change was beneficial only when it moved the participant closer to the requested amplitude. The largest convergence improvements in initial angular error were seen in PD17 (−3.65°), PD31 (−2.71°), PD12 (−2.57°), PD47 (−2.18°), and PD41 (−2.14°). For divergence, the largest improvements were observed in PD30 (−4.38°), PD31 (−2.78°), PD19 (−2.44°), PD12 (−1.54°), and PD13 (−1.16°). However, some participants worsened despite the overall convergence effect, including PD42 for convergence (+1.21°), PD22 (+0.78°), and PD24 (+0.29°), and PD47 for divergence (+1.38°), PD10 (+0.94°), and PD11 (+0.76°).

Other dissociations were also evident. PD10 and PD28 showed marked shortening of convergence latency (approximately 580 ms and 490 ms, respectively), yet divergence endpoint accuracy worsened by 0.32 and 0.44, indicating that faster initiation and better endpoint accuracy were separable. PD12 and PD30 showed large increases in first divergence gain (+0.64 and +0.75) with much smaller and opposite-signed changes in final divergence accuracy (+0.16 and −0.10), suggesting that DBS altered the initial motor response more than the eventual endpoint. Meanwhile, PD17, PD32, and PD41 showed pronounced shifts toward more negative holding in both directions, particularly during convergence, accompanied by some improvement in final accuracy.

Given the marked individual heterogeneity, we asked whether participants could be partitioned into *response subgroups*. Hierarchical clustering was performed in all participants with complete profiles using eight standardized features: on-off changes in final-gain error, first gain, latency, and holding, each measured for convergence and divergence.

A small subgroup consisting of PD29 and PD31 emerged consistently. In the best-performing two-cluster solution, these two participants separated from the remaining with a silhouette value of 0.33, reflecting modest but detectable structure. Both showed large reductions in final-gain error in both directions together with lower first convergence gain, suggesting a similar pattern of DBS-related correction of excessive OFF-state responses. A three-cluster solution additionally identified a possible gaze holding-shift group (PD17, PD24, PD32, PD41, PD48), but separation was weak (silhouette 0.15), and a four-cluster solution mainly isolated PD10 because of marked divergence-latency change (silhouette 0.18). Cluster assignments remained reasonably stable when one participant was removed, but were only moderately robust to omission of individual features (median adjusted Rand index 0.55).

### Computational modeling of pathway activation, volume of tissue activated, and associations with vergence behavior

The heterogeneous effects of STN DBS on vergence may reflect differential engagement of neural circuits governing distinct aspects of binocular motor control. Variability in stimulation location, volume of tissue activated, and recruitment of specific white matter pathways may selectively influence vergence initiation, execution, accuracy, gaze holding, and saccade-assisted strategies. This anatomical variability provides an opportunity to investigate the circuit-level mechanisms underlying these behaviors. Accordingly, we examined associations between modeled activation of STN subregions, peri-subthalamic territories, and white matter pathways and DBS-induced changes in vergence function (Fig 2A,B). Across analyses, each behavioral outcome was defined as the within-participant on-minus-off change. Negative values indicated improvement for latency and absolute-error measures. The top exploratory anatomy–behavior associations are summarized in Table 5, and the multivariable modeling results in Table 6.

**Figure 2:**
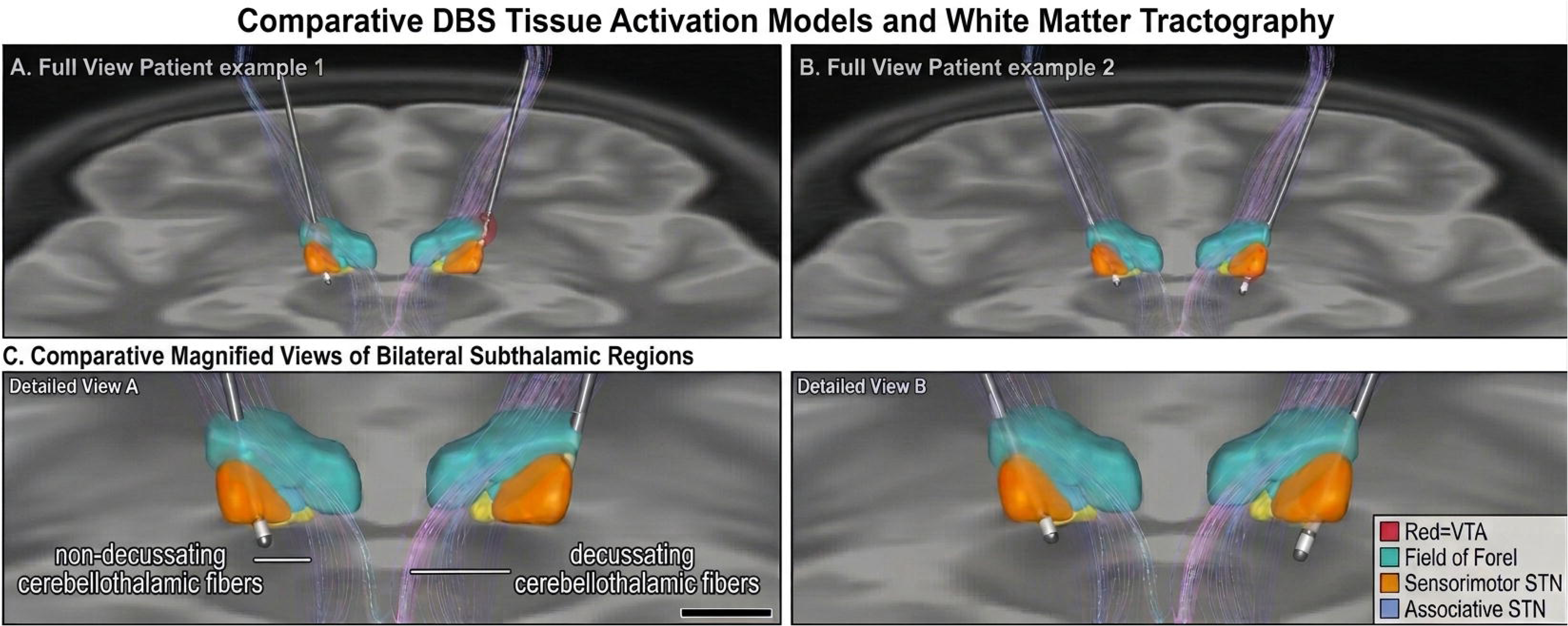
Comparative deep brain stimulation (DBS) tissue activation models and white matter tractography. (A, B) Coronal overview reconstructions of bilateral DBS lead trajectories, volumes of tissue activation (VTA), and overlaid white matter pathways for Patient example 1 (A) and Patient example 2 (B). (C) Magnified views of the bilateral subthalamic regions corresponding to Patient example 1 (Detailed View A) and Patient example 2 (Detailed View B), highlighting the spatial relationship between stimulation fields, target nuclei, and adjacent fiber tracts. Segmented anatomical structures include the Field of Forel (light teal), sensorimotor subthalamic nucleus (STN; orange), and associative STN (light blue, partially obscured beneath the Field of Forel and sensorimotor STN), alongside the VTA surrounding the active electrode contacts (red). Overlaid tractography illustrates non-decussating cerebellothalamic fibers coursing laterally and decussating cerebellothalamic fibers crossing the midline.

**Table 5.**
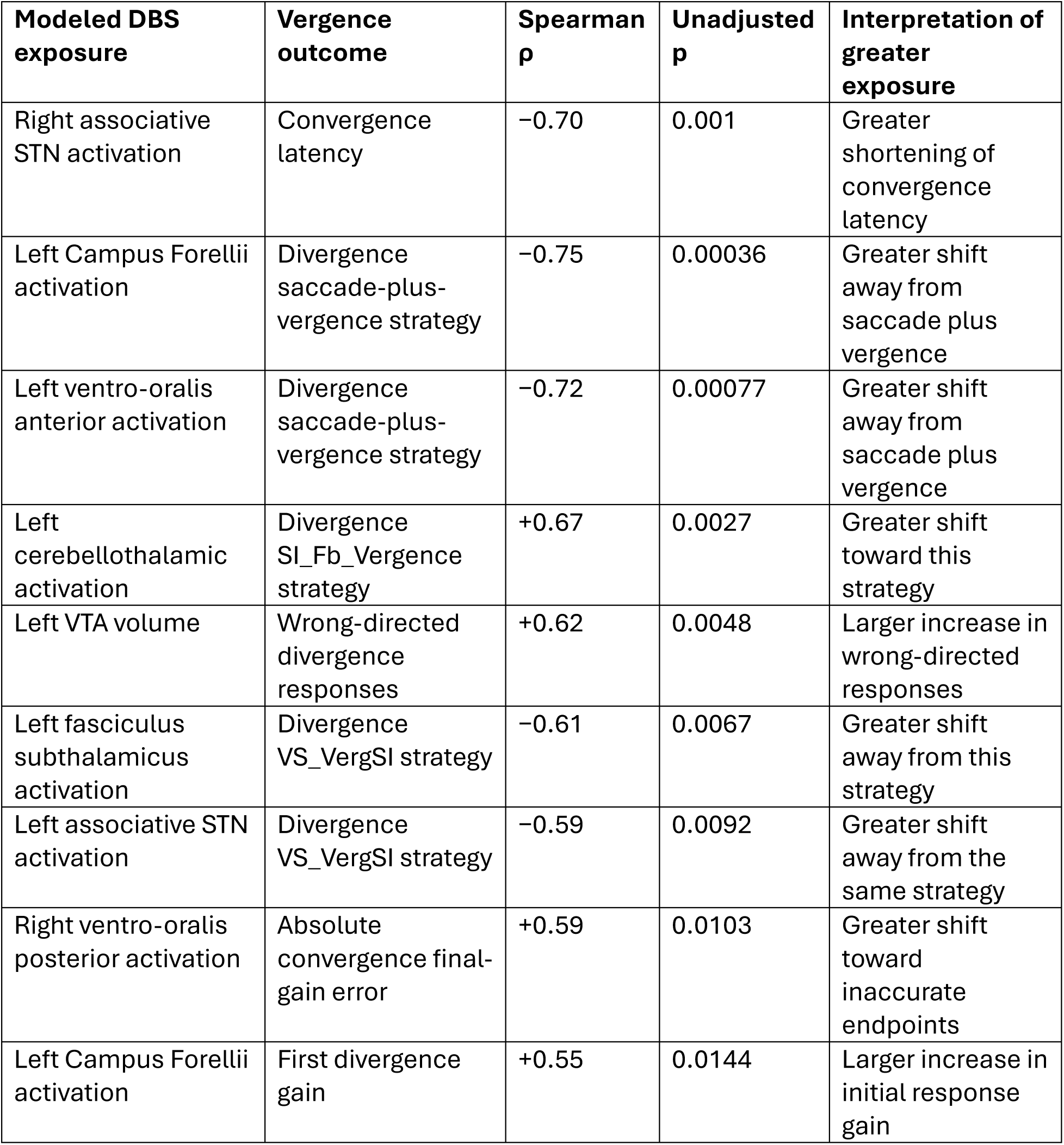
Top exploratory anatomical associations with DBS-related vergence changes.

**Table 6.** Multivariable modeling of anatomical predictors of DBS-related strategy change.

| Outcome | Model / selected predictors | Held-out result vs benchmark | Interpretation |
|---|---|---|---|
| Divergence vergence-plus-saccade strategy | Elastic net: right sensorimotor STN (+), left associative STN (−), left fasciculus subthalamicus (−), right internal capsule (−) | Squared error decreased 16.7%; RMSE 29.2 → 26.6 points | Modest predictive gain, concentrated in a few participants |
| Divergence saccade-plus-vergence strategy | Elastic net / joint strategy model; left Campus Forellii carried a negative coefficient | Squared error decreased 35.0%; RMSE 22.2 → 17.9 points; MAE 17.6 → 14.7 points | Strongest updated strategy-prediction signal, but sensitive to cohort composition |
| Wrong-directed divergence responses | Two-volume model (left and right VTA volumes) | RMSE 18.7 → 15.6 points; squared-error advantage 30% in full cohort, 8% without PD24 | Left VTA volume related to increased wrong-directed responses |
| Divergence saccade-plus-vergence strategy | Two-volume + left fasciculus subthalamicus + left CBT_L model | RMSE 15.3 vs 25.3 for two-volume model and 22.2 for training mean | Exploratory internal improvement that persisted after omission of selected influential participants |
| Convergence latency | Elastic net | RMSE 463 ms (benchmark) vs 465 ms (elastic net) | No predictive improvement |
| Gain, holding, final-gain accuracy | Elastic net | No meaningful predictive advantage | Anatomy did not reliably predict these outcomes |

### Broad anatomical screen identifies candidate associations for convergence latency and divergence strategy

Across the full anatomical screen, the strongest association was observed between greater right associative STN coverage and larger DBS-on shortening of convergence latency (Spearman ρ = −0.70, permutation p = 0.001, n = 19; Table 5). Leave-one-out analyses showed that the correlation with convergence-latency change remained consistently strong (ρ range, −0.78 to −0.65), and the relationship persisted after accounting for total VTA volume (partial rank ρ = −0.62). Greater right associative STN coverage was also associated with both larger right-sided stimulation volumes (ρ = +0.67) and longer OFF-state convergence latency (ρ = +0.58). When both right VTA volume and OFF-state latency were accounted for, the correlation weakened to ρ = −0.43, suggesting that part of the observed effect may reflect greater room for improvement in participants who were slower at baseline.

This association was notably weaker for divergence latency, and a direct comparison using each participant’s convergence-change minus divergence-change score also yielded only a weak relationship (Table 5).

Although group-level strategy frequency changed little (Table 3), individual divergence strategy changes were associated with modeled left-sided stimulation anatomy (Table 5). Greater overlap with the left Field of Forel (ρ = −0.75, unadjusted p = 0.00036) and left ventro-oralis anterior (ρ = −0.72, unadjusted p = 0.00077) accompanied larger reductions in combined saccade-plus-vergence responses.

### Associations with endpoint error

Greater stimulation overlap with the right ventro-oralis posterior was associated with larger convergence endpoint errors with DBS ON, both in degrees (ρ = 0.61, p = 0.0073) and relative to target demand (absolute final-gain error, ρ = 0.59, p = 0.0096). Greater overlap with the right nucleus reticulatus polaris showed a similar pattern for divergence (angular error, ρ = 0.58, p = 0.012, and absolute final-gain error, ρ = 0.53, p = 0.021) (Table 5). Associations with initial angular error were weaker and did not reach nominal significance. Total VTA volume showed weaker associations than regional overlap, with its largest correlations observed for the two divergence endpoint error measures. None of the accuracy associations survived correction for multiple comparisons (minimum q = 0.647).

### First vergence gain showed weak and exploratory anatomical associations

Greater left Field of Forel overlap showed the strongest association with a DBS-related increase in signed first gain for divergence (ρ = +0.55, p = 0.015, n = 19; Table 5). The correlation was stable in leave-one-out analyses (ρ = +0.51 to +0.63) but did not survive correction for multiple comparisons (q = 0.93). Neither modeled fiber activation nor total stimulation volume showed a clear association with first gain in either direction.

Greater overlap was only weakly associated with a reduction in absolute initial error (|first gain − 1|, ρ = −0.22, p = 0.372) and correlated with lower off-state first divergence gain (ρ = −0.55). This exploratory finding therefore points to a larger initial divergence response with DBS, potentially dependent on baseline gain, with little evidence of improved initial accuracy.

### Multivariable prediction was strongest for divergence strategy

We used elastic-net models to test whether combinations of anatomical features improved prediction beyond individual associations. The 42 candidate predictors included hemisphere-specific fiber activation, regional stimulation coverage, and left and right VTA. In nested cross-validation, we predicted each participant’s DBS on–off change from models trained on the remaining participants. The mean change in the training set served as the benchmark (Table 6).

Prediction improved most for divergence strategy, particularly the proportion of trials combining saccades and vergence. Greater left Field of Forel coverage was a leading correlate of reduced use of this strategy. Anatomical models reduced squared prediction error by 26.7–35.0% relative to the benchmark, although the advantage diminished substantially after influential participants were excluded. Under the original strategy classification, greater right sensorimotor STN coverage predicted more frequent use of the vergence-plus-saccade category, whereas greater left associative STN coverage, left fasciculus subthalamicus activation, and right internal capsule coverage each predicted less frequent use. All four predictors were selected in 18 of 19 training sets, with modest improvement in prediction for held-out participants.

In exploratory regression, each 1-SD increase in left VTA volume was associated with an 11.1-percentage-point increase in wrong-directed divergence responses (95% CI, +1.5 to +20.6), although statistical evidence for the bilateral VTA model as a whole was limited (p = 0.061). Adding left fasciculus subthalamicus and left cerebellothalamic pathway activation to bilateral VTA volumes improved prediction of changes in combined saccade-plus-vergence responses. Root mean squared error (RMSE) was 15.3 percentage points with the pathways included, compared with 25.3 for volumes alone and 22.2 for the training-mean benchmark. This advantage persisted when influential participants were excluded one at a time, suggesting that pathway engagement added predictive information beyond stimulation volume within this cohort. Because strategy proportions sum to 100%, a decrease in one category corresponds to an increase in one or more others.

Elastic-net models did not meaningfully improve prediction of latency, gain, accuracy, or gaze holding. Despite the strong univariable association with right associative STN coverage, convergence latency RMSE was 465 ms versus 463 ms for the benchmark. For divergence latency, squared prediction error increased by 13.0%. Squared error for final divergence gain decreased by only 0.15%, while prediction of absolute final-gain error worsened in both directions. For gaze holding, squared error increased by 33.4% for convergence and 3.8% for divergence.

Elastic net improved none of the six accuracy outcomes: initial angular error, endpoint angular error, and absolute final-gain error for each direction. In separate regressions accounting for DBS off-state error, adding bilateral VTA volumes yielded only a small improvement for divergence absolute final-gain error (RMSE, 0.401 to 0.394). Adding left fasciculus subthalamicus and left cerebellothalamic pathway activation increased held-out prediction error for all six accuracy outcomes compared with using OFF-state error alone (Table 6).

### Anatomical models explained effects on gaze-holding

The behavioral analyses showed a clear average DBS ON–OFF effect on gaze holding in convergence or divergence position. Gaze holding variance decreased with DBS ON in 12 out of 19 participants (Group 1) and remained unchanged or increased in seven (Group 2). Group assignment was based on the ON-minus-OFF change in holding variance, with convergence and divergence weighted equally within each participant. For each anatomical measure, we averaged the left and right percentages. The cumulative score summed the bilateral averages for Field of Forel coverage, cerebellothalamic pathway activation. The specific rationale for choosing these pathways was that its coverage with VTA would retrogradely modulate the cerebellar outflow, influencing the fidelity of vergence neural integrator, affecting gaze holding in convergence or divergence state.

The median cumulative stimulation score was 13.48 in Group 1 and 9.31 in Group 2 (rank-biserial effect size = 0.48, exact permutation *p* = 0.100, adjusted *q* = 0.201). Median exposure was numerically higher in Group 1 for all three components. Bilateral cerebellothalamic pathway activation showed the largest rank separation between groups (rank-biserial effect size = 0.49, *p* = 0.071, *q* = 0.201). All 19 participants had some modeled Field of Forel coverage on at least one side. Across participants, the correlation between the cumulative score and the magnitude of change in holding variance was weak (Spearman ρ = −0.23, *p* = 0.351). Thus, participants with improved holding had higher median cumulative exposure, but the group difference was statistically inconclusive.

### Stimulation anatomy distinguishes exploratory patterns of gaze holding and vergence initiation

To examine whether stimulation anatomy distinguished patterns of gaze stability, we grouped ON– OFF changes in gaze-holding variance together with cumulative exposure of field of Forel and cerebellothalamic projections. Convergence and divergence observations were included together, with separate analyses for each hemisphere (Fig 3A,B; Table 7). The higher right-sided exposure cluster showed reduced gaze-holding variance, whereas the lower-exposure cluster showed little average change. Mean cumulative exposures were 29.67 and 4.38, with corresponding variance changes of −0.00513 and +0.00074 deg². The between-cluster difference was −0.00586 deg² (95% CI, −0.00870 to −0.00302; nominal *p* = .00040; BH-adjusted *q* = .00079). A similar pattern occurred for left-sided stimulation. Mean cumulative exposures were 86.74 and 13.51, with variance changes of −0.00462 and −0.00141 deg², respectively (difference, −0.00321 deg²; 95% CI, −0.00577 to −0.00065; nominal *p* = .017; *q* = .017). Thus, greater cumulative exposure characterized clusters with steadier post-vergence gaze holding.

**Figure 3:**
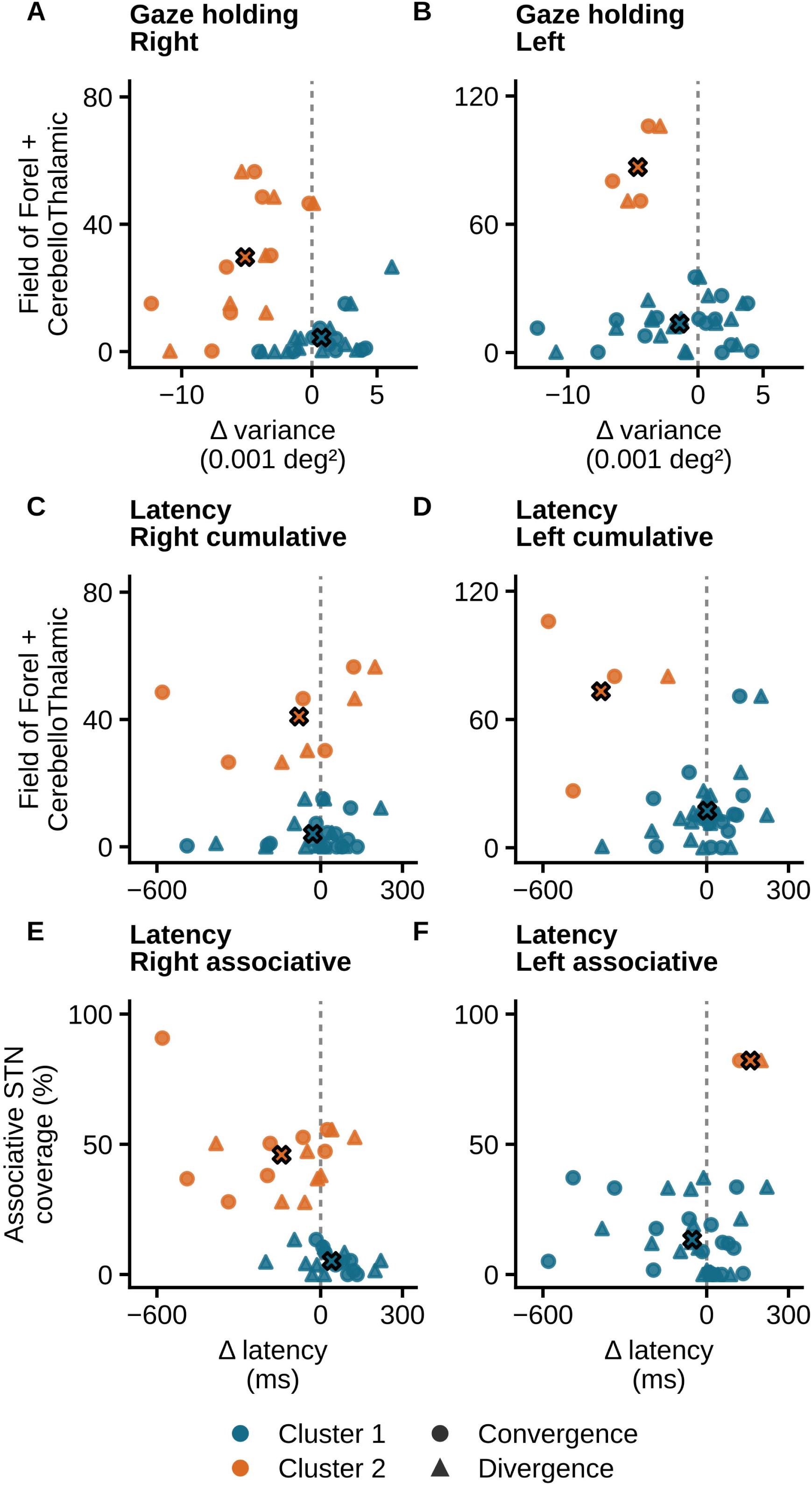
Clusters of gaze-holding and latency changes in relation to modeled DBS exposure. Panels A and B show gaze-holding variance change versus right and left cumulative Field of Forel + Cerebellothalamic pathway coverage. Panels C and D show latency change versus the corresponding right and left cumulative exposure. Panels E and F show latency change versus right and left associative STN coverage. Circles indicate convergence and triangles divergence. Blue and orange indicate clusters 1 and 2, respectively; X symbols mark cluster means. Separate two-cluster k-means models were fitted for each panel after standardizing both coordinates. Cluster 1 has lower mean exposure. Negative ON minus OFF changes indicate steadier gaze holding or faster initiation. Gaze-holding variance is plotted in 10⁻³ deg²; statistics retain deg² units. Cumulative scores combine regional coverage and fiber activation percentages with different denominators and do not measure unique tissue volume.

**Table 7.** Selected associations between modeled stimulation exposure and DBS on-minus-off clinical change.

| **Clinical outcome** | **Stimulation measure** | **Spearman ρ** | **Unadjusted *p*** | **Partial rank ρ** |
|---|---|---|---|---|
| Motor score | Right cerebellothalamic activation | −0.52 | 0.025 | −0.35 |
| Motor score | Left ventro-oralis posterior activation | +0.56 | 0.014 | +0.49 |
| RAF ruler distance | Right frontal corticopontine tract activation | +0.55 | 0.045 | +0.43 |
| Distance to pen | Right substantia nigra activation, combined pars compacta/pars reticulata activation | −0.66 | 0.0050 | −0.67 |
| Distance to pen | Left cerebellothalamic pathway activation | −0.53 | 0.033 | −0.57 |
| Distance to pen | Left VTA volume | −0.43 | 0.090 | — |

We next examined whether cumulative exposure and associative STN coverage distinguished changes in movement initiation (Fig 3C-F; Table 7). The higher left cumulative-exposure cluster showed a mean latency reduction of 387 ms, compared with a 2-ms increase in the lower-exposure cluster (difference, −389 ms; 95% CI, −597 to −181; nominal *p* = 0.00098; *q* = 0.0020). The corresponding right-sided comparison showed no clear latency difference (−50 ms; 95% CI, −258 to +158; *p* = 0.617). Greater right associative STN coverage characterized a cluster with shorter latency during DBS on: latency decreased by 143 ms in the higher-coverage cluster and increased by 40 ms in the lower-coverage cluster (difference, −183 ms; 95% CI, −307 to −60; nominal *p* = 0.0060; *q* = 0.0072). Left associative STN comparisons were not estimated because the higher-coverage cluster contained only one participant. Comparisons accounted for repeated observations within participants. Because clustering used the same outcomes subsequently compared, and exclusions depended on response values, these statistics describe the selected clusters and do not independently confirm stimulation–behavior associations.

### Associations of stimulation anatomy with clinically measured motor and ophthalmological responses

Of the 24 participants in the paired DBS vergence cohort, 19 had imaging and paired motor scores, 14 had paired RAF ruler measurements, and 17 had paired distance-to-pen measurements. Clinical changes were calculated as DBS on-minus-off. Negative changes indicate lower motor scores or shorter convergence distances with stimulation. Associations with modeled fiber activation, regional overlap of the VTA, and overall VTA volume were evaluated using Spearman rank correlations and two-sided permutation tests.

Greater right cerebellothalamic activation was associated with a larger reduction in motor score with DBS on (ρ = −0.52, unadjusted *p* = 0.025). The right reticular tract measure yielded the same rank correlation. Greater left ventro-oralis posterior overlap was associated with a smaller reduction or an increase in motor score (ρ = +0.56, *p* = 0.014). After adjustment for off-state motor score and total VTA volume, the partial rank correlations were −0.35 for right cerebellothalamic pathway activation and +0.49 for left ventro-oralis posterior overlap. Overall VTA volumes showed little association with motor score change (|ρ| ≤ 0.11, all *p* ≥ 0.662).

Greater right frontal corticopontine tract activation was associated with a longer RAF ruler distance (ρ = +0.55, unadjusted *p* = .045). After adjustment for OFF-state RAF ruler distance and total VTA volume, the partial rank correlations were +0.62 and +0.43, respectively. Overall VTA volumes showed weak associations with RAF ruler distance change (|ρ| ≤ 0.19, all *p* ≥ .501).

For distance to pen, greater overlap with the right substantia nigra region, defined by the combined pars compacta/pars reticulata atlas label, accompanied a larger shortening with DBS ON (ρ = −0.66, unadjusted *p* = 0.0050). Greater activation of the left corticospinal tract (ρ = −0.55, *p* = .025) and left cerebellothalamic pathway (ρ = −0.53, *p* = 0.033) also accompanied shorter distances. After adjustment for OFF-state distance to pen and total VTA volume, the corresponding partial rank correlations were −0.67, −0.60, and −0.57. Among the overall volume measures, left VTA volume showed the largest correlation with distance to pen (ρ = −0.43, *p* = 0.090).

## Discussion

Changing viewing depth requires the brain to establish a new binocular alignment and sustain it after the movement ends. Our findings suggest that STN-region DBS can affect these components differently. Its clearest effect was on post-vergence gaze stability: eye-position variance decreased by approximately one-third across convergence and divergence. The initial convergence response also landed closer to the target, whereas average changes in signed gain, final accuracy, latency, and movement strategy were less consistent. This pattern suggests that a subcortical perturbation can improve the maintenance of binocular alignment with relatively little average change in how the eyes attain it. Exploratory anatomical analyses identified candidate contributions from associative STN territories and the fields of Forel with adjacent cerebellothalamic projections. These observations raise the possibility that stimulation influences movement preparation and saccade–vergence coordination through basal ganglia circuits, while modulation of cerebellar signals reaching midbrain vergence circuitry helps stabilize the attained eye position. The reduction in post-vergence variability provides the strongest behavioral basis for examining this proposed distinction.

### Maintaining alignment after a depth shift

STN-region DBS reduced vergence holding variance after both convergence and divergence, with little evidence of a change in final gain. This finding is consistent with improved fidelity of vergence integration. Once the eyes reached a new vergence angle, they fluctuated less around it, even though the average achieved angle changed little. Participants whose gaze holding improved had greater cumulative modeled coverage of the fields of Forel and cerebellothalamic fibers than those whose gaze holding was unchanged. Together, these observations support the hypothesis that retrograde modulation of cerebellar outflow by engaging field of Forel and cerebellothalamic projections would improve the fidelity of vergence integration, expressed behaviorally as a steadier gaze holding after the vergence movement ends.

One possibility is that stimulation of cerebellothalamic axons alters the cerebellar signals that tune midbrain vergence circuitry. Field of Forel and cerebellothalamic track modulation can antidromically modulate deep cerebellar nuclei(Tolbert et al., 1978), and modulation of the fastigial and posterior interposed outputs reach the SOA and adjacent central mesencephalic reticular formation(Bohlen et al., 2021). Although the axons recruited by DBS in our participants belonged to neurons that also influence these midbrain regions remains a speculation, the combined coverage measure identifies a region in which engagement of neighboring or overlapping pathways are associated with better gaze holding. The variance in gaze holding also reflects both the stability of the tonic position command and the effectiveness of visual disparity feedback(Quinet et al., 2025, Semmlow et al., 1993, Semmlow et al., 2019).

### Initiating a depth shift

The association between greater right associative STN coverage and shorter convergence latency is compatible with modulation of movement preparation or release. Prefrontal inputs to the associative STN provide an anatomical basis for such an effect(Haynes and Haber, 2013). A candidate downstream route runs through the substantia nigra pars reticulata and superior colliculus to midbrain vergence circuitry. Primate experiments link nigral output to the release of saccades, and the rostral superior colliculus carries slow vergence signals and projects to the supraoculomotor area(Hikosaka and Wurtz, 1985, May et al., 2021, Van Horn et al., 2013). Stimulation might shorten the time required to disengage fixation, select a response to disparity, or release its first detectable motor component. Latency alone leaves these possibilities open.

### Coordinating saccades and vergence

Combined saccade-plus-vergence responses predominated in both stimulation states. Their persistence is consistent with the physiology of three-dimensional gaze shifts, in which saccades transiently facilitate vergence and influence how rapidly the eyes reduce disparity(Busettini and Mays, 2005). The strongest anatomical signal for those individual differences was reduced use of combined responses during divergence with greater left Field of Forel overlap. Left ventro-oralis anterior overlap and left fasciculus subthalamicus recruitment showed associations in the same direction. The proximity of pallidothalamic and cerebellothalamic fibers in this region makes Field of Forel overlap a marker of exposure to the border region(Gallay et al., 2008). It leaves open contributions from pallidothalamic output, pallidal–subthalamic interactions, and adjacent cerebellothalamic fibers.

The modeled stimulation overlap with the campi Foreli, pallidothalamic projections, and cerebellothalamic projections suggest a complementary route to modulate saccade vergence collaboration to achieve gaze shift in depth. Within this anatomy, pallidal output could bias the anticipated movement strategy, including whether a smooth response is likely to suffice and when a saccade should be released if it is recruited. Cerebellar output could calibrate the selected response, including how much of the remaining vergence error the saccade should remove and how the eyes settle afterward(Busettini and Mays, 2005, Ilinsky et al., 2018, Sakai et al., 1996, Takeichi et al., 2005).

Cerebellar output, caudal fastigial and posterior interposed nuclei project to distinct parts of the midbrain cMRF(Bohlen et al., 2021, May et al., 1990, May et al., 1992); the region relevant to saccades. Saccade–vergence burst neurons in this region discharge in relation to vergence velocity; the rapid vergence contribution depends on the interaction of a saccadic burst with the vergence error remaining when the saccade begins(Busettini and Mays, 2005, Quinet et al., 2020). In this way STN DBS, via recruiting and antidromically modulating cerebellothalamic projection can modulate when the saccadic burst arrives relative to the evolving vergence error, how strongly the burst contributes to the change in binocular alignment(Gallay et al., 2008, McIntyre et al., 2004).

### Implications for sensorimotor control

Binocular near viewing depends on how the brain uses visual cues to align and focus the eyes and then keep them steady. Our findings suggest that STN DBS may influence different parts of this process. Video-oculography showed steadier gaze after a depth shift, with greater combined stimulation coverage of the field of Forel and adjacent cerebellothalamic pathways among participants who improved. The bedside tests probed how well binocular alignment or single vision was maintained as disparity, blur, and proximity cues changed together. Greater left cerebellothalamic activation was associated with a closer pen-tip breakpoint, whereas greater right frontal corticopontine activation and internal capsule overlap were associated with a more distant RAF ruler breakpoint. These exploratory findings raise the possibility that cerebellar outflow helps stabilize an attained vergence position, while frontal signals relayed through the pons and cerebellum may shape how visual cues drive the midbrain near response(Bohlen et al., 2021, Gallay et al., 2008, Gamlin and Clarke, 1995, Gamlin and Yoon, 2000, Horwood and Riddell, 2009, Zhang and Gamlin, 1998). The bedside associations did not survive correction for multiple comparisons. A prospective within-participant study pairing targeted stimulation with eye tracking, bedside testing, and reading measures could determine whether steadier gaze also helps patients with Parkinson’s disease maintain single vision as viewing demands change.

## Data Availability

All data produced in the present study are available upon reasonable request to the authors

## Authors’ Roles

IQ: Data collection, Data analysis, Manuscript writing, Manuscript editing

AG: Data analysis, Manuscript editing

CK: Data collection, Manuscript editing

FG: Conceptualizing project, Data collection, Data analysis, Manuscript editing

AG: Conceptualizing project, Data collection, Data analysis, Manuscript writing and editing

